# Clinician-Centered Evaluation of Large Language Model-Generated Discharge Summaries for Longer Hospitalizations: Insights from Hospitalists and Primary Care Physicians

**DOI:** 10.64898/2026.06.03.26354858

**Authors:** Tyler Osborne, Tauhid Mahmud, Xia Zheng, Sai Jampala, Sadia Abbasi, Stephanie Hong, Kimberly Kranz, Susan Lee, Patricia Ng, Kerim Odekon, Lindsey Schachter, Robert Sexton, Tracey Spinnato, Mathew Tharakan, Zhi Wu, Fusheng Wang, Rachel Wong

**Affiliations:** Stony Brook University, Department of Computer Science, Stony Brook, NY; Stony Brook University Department of Biomedical Informatics, Stony Brook, NY; Stony Brook University, School of Communication and Journalism; The Alan Alda Center for Communicating Science, Stony Brook, NY; Stony Brook Medicine, Department of Internal Medicine, Stony Brook, NY

## Abstract

Although large language models (LLMs) have shown promise for discharge summary generation, their value may be greater in longer hospitalizations, where increasing documentation volume and complexity increase both clinician burden and the risk of communication failures during transitions of care. Prior evaluations of LLM-generated discharge summaries have largely involved shorter stays and have rarely examined receiving-clinician priorities or incidental finding reporting. We compared LLM-generated and human-authored discharge summaries for 60 Internal Medicine hospitalizations lasting 7 to 21 days, with paired assessment by hospitalists and primary care physicians (PCPs). Clinician reviewers preferred LLM-generated summaries for 95% of encounters and rated them higher for quality, readability, factuality and completeness. PCPs, the primary recipients responsible for post-discharge care, found that LLM-generated summaries were better for understanding and communicating hospital care to patients, and providing follow-up care. LLM-generated summaries had fewer annotated errors, primarily due to fewer omissions, without increased estimated harm potential or likelihood compared with human-authored summaries. Benefits of LLM-generated summaries were especially salient for PCPs, who identified more omissions with greater downstream likelihood of harm than hospitalists. This underscores the importance of designing transition documents around the needs of clinicians assuming care post-discharge. LLM identification of radiology incidental findings was generally accurate and appropriate, suggesting potential to improve follow-up of clinically relevant findings. These findings extend prior work by demonstrating clinical value of LLMs in summarizing longer, complex hospitalizations and highlighting the value of stakeholder-centered design in clinical AI systems. Together, they support supervised LLM-assisted discharge summarization as a tool to reduce cognitive burden, improve documentation quality, and enhance transition-of-care communication.

## Introduction

Hospital discharge summaries are a critical component of safe transitions of care because they communicate the hospitalization course, treatment decisions, medication changes, unresolved issues, and follow-up needs to outpatient clinicians responsible for ongoing care. High-quality discharge summaries are important for patient safety and continuity of care, as incomplete or unclear documentation can contribute to medication errors, missed follow-up, and fragmented care after discharge^(1–4)^. However, writing effective discharge summaries is burdensome, particularly for longer hospitalizations, where clinicians must synthesize extensive and often complex clinical information in a concise transition-of-care document^(5,6)^.

Evidence on the use of large language models (LLMs) for discharge summary documentation is growing rapidly, with multiple retrospective studies demonstrating that LLM-generated summaries achieve comparable overall quality to physician-written summaries. Williams et al. (2025) found that LLM-generated summaries outperformed physicians on conciseness and coherence, with no significant difference in overall quality; blinded reviewers showed no preference for one over the other^(7)^. Mehri et al. (2026) found no difference in total quality scores between LLM-generated and human-authored discharge summaries^(8)^. Challener et al. (2026) found that LLM-generated summaries outperformed clinicians across nine quality domains, with the greatest advantage in comprehensiveness^(9)^. A recent prospective study by Grolleau et al. (2026), which incorporated an agentic LLM workflow into discharge operations at a medical center, provides even stronger support for the real-world use of LLM-generated discharge summaries. They found that physicians incorporated LLM-generated content into 57% of their final discharge notes, and that among feedback collected on 100 summaries, 88% were rated as having no harm potential. Importantly, physician burnout scores decreased despite insignificant time savings, suggesting that LLMs may reduce provider burden through cognitive offloading^(10)^.

However, important gaps remain in the current literature on LLM-generated discharge summaries. First, prior studies have largely focused on shorter hospitalizations and have not evaluated longer stays, likely because of the substantial effort required for manual chart review and the technical challenges posed by LLM context window limitations. Longer hospitalizations represent a particular challenge because they often generate large volumes of fragmented, repetitive, and clinically complex documentation, making discharge summarization substantially more burdensome for clinicians than shorter admissions^(10)^. Clinicians may therefore derive the greatest benefit from systems capable of maintaining summary quality despite increasing documentation burden. Addressing this challenge also requires technical approaches that enable processing of electronic health records (EHRs) extending beyond current model context window limitations.

Second, although prior studies have shown that LLM-generated discharge summaries can achieve quality and readability comparable to or better than human-authored summaries,^(9,11)^ there is limited work on how these summaries are perceived by different stakeholders in transition of care. Only one study stratified evaluations by reviewer type, and they found that primary care physicians (PCPs) and nursing home physicians rated LLM-generated summaries as significantly more concise, whereas hospitalists did not demonstrate the same preference^(12)^. More broadly, prior literature demonstrates important differences between hospitalist and PCP priorities for discharge summaries that are highly relevant to LLM design. PCPs consistently identify diagnoses, medication changes, pending tests, and follow-up plans as critical for outpatient care, yet these elements are frequently incomplete or missing from discharge summaries in routine practice.^(2,13)^ Chatterton et al. found that PCPs prefer concise summaries focused on next steps, including clearly flagged incidental findings, explicit rationale for medication changes, and specific follow-up instructions, while excessive length was a source of frustration^(14)^. Although inpatient and outpatient clinicians generally agree on the core components of discharge summaries, studies suggest that hospitalists tend to prioritize detailed descriptions of the hospitalization whereas PCPs place higher priority on actionable follow-up information such as discharge medications.^(15)^ Together, these findings suggest that LLM-generated discharge summaries may benefit from prompting strategies specifically designed around PCP workflow needs, such as formatting around action items and explicit highlighting of medications results, pending results and incidental findings.

Third, prior work has not evaluated LLM-based extraction of incidental findings from radiology reports in the context of discharge summary generation, which is important given the risk of missed follow-up and patient harm. Incidental findings are common and frequently lack appropriate follow-up, yet determining which findings are clinically actionable for outpatient care requires nuanced clinical judgment.^(16,17)^ Although prior studies have shown that LLMs can identify incidental findings from radiology reports with reasonable accuracy,^(18–20)^ it remains unclear how well these systems perform when integrated into discharge summaries, where excessive reporting may also contribute to information overload for outpatient clinicians.

To address these gaps, we conducted a clinician-centered evaluation of LLM-generated discharge summaries for longer hospitalizations (7–21 days) and compared them with human-authored summaries for overall performance and documentation errors. We developed a scalable summarization pipeline capable of processing EHR documents that exceed current LLM context-window limits and incorporated a workflow for identifying and reporting radiology incidental findings. To assess stakeholder-specific perspectives, we designed prompts around PCP informational priorities and engaged both hospitalists and PCPs in role-specific, and workflow-aligned evaluations. By combining technical innovation with stakeholder-centered evaluation, this study provides new evidence on the clinical utility, safety, and workflow implications of LLM-generated discharge summaries for supporting transitions of care.

## Methods

### Study Cohort

We extracted clinical notes, imaging reports, and discharge summaries from 400 historical inpatient encounters on the Internal Medicine service at Stony Brook University Hospital between January 1, 2023 and December 31, 2024, with lengths of stay ranging from 7 to 21 days. Encounters without discharge summaries or with fewer than 25 clinical notes were excluded. For each encounter, we extracted all clinical notes from the EHR, which included 654 unique note types across multiple categories, including emergency department notes, consult notes, admission and history and physical notes, progress notes, care management notes, addenda, and event notes. Note types were reviewed by a physician informaticist and filtered for inclusion as LLM inputs based on clinical relevance for discharge summary generation. All data storage and computation was performed via a secure shell on an in-network computing cluster. Inputs to GPT-5.2 were submitted through an API to a private HIPAA-compliant Microsoft Azure OpenAI Service instance. The study was approved by the Stony Brook University Office of Research Compliance.

### Encounter Processing

Notes for each encounter extracted from the Oracle Cerner EHR were provided in rich text format (RTF) and converted to plain text for LLM processing using the Python striprtf library (Python 3.12). Plain-text notes and encounter-level metadata were stored in a SQLite database using sqlite3 (Python 3.12). Notes were ordered chronologically, and those published after the final progress note for the encounter were excluded to prevent leakage of information unavailable at the time of discharge. Notes containing fewer than 40 characters were also removed, as these were primarily attestation entries. For each encounter, the historical human-authored discharge summaries were extracted for comparison with the LLM-generated summaries.

### LLM Discharge Summary Generation

A flow diagram of the technical steps in LLM discharge summary generation is shown in Supplementary Figure S1. Because GPT-5.2 has a context limit and longer encounters could not be processed in a single pass, we divided the clinical documents into 50,000-token segments and inputs were assembled in a SQLite database. The LLM was prompted to create a chronological, bulleted draft of key components of the discharge summary (Supplementary Figure S2) using iterative refinement with a sliding window over the note segments. The model generated an initial summary from the first segment of the record and then updated the summary as successive segments were processed, ensuring that each call remained within the window limit. We used a window size of 50,000 tokens to balance token costs, latency and output quality. In parallel, a separate prompt was developed for identification and extraction of incidental findings from radiology reports (Supplementary Figure S3). The LLM was then prompted to consolidate the chronological, bulleted draft and incidental findings into a final narrative summary (Supplementary Figure S4). Pipeline development utilized LangChain and LangGraph in Python 3.12.

### Discharge Summary Evaluation

Evaluation of both human-authored and LLM-generated discharge summaries was conducted with reviews by 12 attending physicians (6 hospitalists and 6 PCPs with representation from inpatient, outpatient, and informatics leadership) using quantitative evaluation followed by focus group debriefing sessions. A stratified random sample of 60 encounters was selected to match the length of stay distribution of the full dataset. Each physician was assigned to review 10 encounters, with every encounter independently evaluated by one hospitalist and one PCP. Reviews were conducted via a custom web application deployed behind the University Hospital firewall; Supplementary Figure S5 shows example screenshots demonstrating the main features of the web application. Evaluators were provided access to both human-authored and LLM-generated discharge summaries, all clinical notes that were used as inputs, and keyword search functionality for locating relevant information within the EHR documentation. The user interface also included features for completing survey questions, annotating and scoring errors within the human-authored and LLM-generated discharge summaries, and a separate section for evaluation of incidental finding reporting.

The evaluation for all reviewers included four questions with a 5-point Likert scale to assess overall quality, conciseness/readability, factuality and completeness^(11)^ as well as preference for the human-authored versus LLM-generated summary. PCPs were assigned three additional survey questions on ease of understanding, verbalizing to patients and caregivers, and following up relevant aspects of the hospitalization (Supplementary Table S1). To evaluate discharge summary errors, reviewers were instructed to identify and annotate errors by type (inaccuracies and omissions) for both human-authored and LLM-generated discharge summaries (Supplementary Table S2). For each error annotation, reviewers provided a brief description and rated the likelihood and potential for patient harm using an adapted Agency for Healthcare Research and Quality harm scale (Table 1).^(7,12)^ For encounters where the LLM detected clinically relevant incidental findings, reviewers were presented with the finding and asked to rate whether it was reported accurately and whether they thought reporting was appropriate (Supplementary Table S3).

**Table 1.** Scores and definitions of potential and likelihood of harm included in the instructions for physician reviewer error annotation.

| Harm Scores | Harm Potential Definition | Harm Likelihood Definition |
| --- | --- | --- |
| 0 (None) | No potential for harm | Impossible (0 in 10M) |
| 1 (Minimal) | Potential for emotional distress or inconvenience (mild and transient anxiety or pain or physical discomfort) | Nearly Impossible ($\leq 1$ in 10M) |
| 2 (Minor) | Potential for requiring additional treatment | Extremely Rare (1 in 1M) |
| 3 (Moderate) | Potential for temporary harm (bodily or psychological injury, but likely not permanent) | Very Rare (1 in 100K) |
| 4 (Significant) | Potential for permanent harm (lifelong bodily or psychological injury or increased susceptibility to disease) | Rare (1 in 10K) |
| 5 (Serious) | Potential for lifelong bodily or psychological injury or disfigurement | Possible (1 in 1,000) |
| 6 (Severe) | Potential for severe permanent harm | Likely (1 in 100) |
| 7 (Catastrophic) | Potential for death | Near Certain ($>1$ in 10) |

To align reviews with real-world clinical workflows, hospitalists were instructed to review all inpatient clinical notes prior to evaluation of the human-authored and LLM-generated discharge summaries, and PCPs were instructed to first review the discharge summaries and then corroborate findings with the inpatient clinical notes (Supplementary Figure S6). Reviewer training was conducted via virtual meetings prior to evaluation. In addition, three virtual, post-review, semi-structured debriefing sessions were held with physician evaluators to discuss findings from the surveys and error annotations and to obtain qualitative feedback on their perspectives. Sessions were recorded and transcribed verbatim. We analyzed the resulting transcripts with a hybrid approach that combined human coding with LLM assistance. Supplementary Table S4 summarizes the themes that emerged during the debriefing sessions.

### Statistics and Reproducibility

A one-sided paired Wilcoxon signed rank test was used to compare physician reviewer Likert-scale ratings of LLM-generated and human-authored discharge summaries, testing whether LLM-generated summaries were rated more favorably. One-sided testing was also used to compare the differences in ratings between hospitalists and PCPs to evaluate whether PCPs perceived a greater gap in performance than hospitalists between LLM-generated and human-authored summaries. Error annotation counts by provider type and error type were compared using two-sided Wilcoxon signed rank testing. Two-sided Mann-Whitney U tests were used to compare the harm potential and harm likelihood score distributions for errors with human-authored versus LLM-generated summaries and by provider type. Associations between length of stay and mean Likert rating were assessed using two-sided Spearman rank correlation tests, separately for human-authored and LLM-generated summaries. Nonparametric tests were used because Likert ratings and annotation counts were not assumed to be normally distributed. For all tests, p < 0.05 was considered statistically significant.

### Data and Code Availability

The underlying clinical notes used to generate the discharge summaries evaluated in this study are private health information and are therefore unable to be released to the public. However, we release the code and raw, de-identified results used to generate statistics at this link: https://github.com/StonyBrookDB/Clinician-Centered-LLM-Discharge-Summaries

## Results

Of the 400 Internal Medicine inpatient encounters extracted, 385 met inclusion criteria; 42% (162/385) were female and ages ranged from 19 to 101 years (mean 72, SD +/- 16). A random sample of 60 encounters was selected, stratified to match the length of stay distribution of all the encounters. 370 of the original 654 note types were retained as LLM inputs, yielding an average reduction of 55% in EHR text volume per encounter. Length of stay ranged from 7 to 21 days along the distribution shown in Supplementary Table S5. Length of stay values over 14 were intentionally oversampled to better represent longer stays in the evaluation.

### Evaluation of LLM-Generated vs. Human-Authored Summary Performance and Errors Quantitative Findings

For most encounters (95%), the physician reviewers preferred the LLM-generated summary to the human-authored summary. The mean scores on the physician reviewer surveys were higher for LLM-generated than human-authored discharge summaries across all metrics: quality (mean [SD] 4.71 [0.47] vs 3.16 [1.08]; p = 1.40e-17), readability (4.66 [0.56] vs 3.39 [1.04], p = 3.25e-15), factuality (4.62 [0.62] vs 3.73 [0.85], p = 1.08e-13), and completeness (4.69 [0.53] vs 3.42 [1.11], p = 2.67e-15). PCP reviewers rated LLM-generated summaries as easier to understand than human-authored summaries (4.75 [0.44] vs 3.20 [1.12], p = 7.40e-10), easier to communicate to patients and caregivers (4.75 [0.47] vs 3.27 [1.12], p = 2.09e-9), and easier to use for providing follow-up care (4.68 [0.57] vs 3.20 [1.19], p = 4.37e-9) than the human-authored summaries. The mean Likert scores for physician reviewer survey questions are shown in Table 2.

**Table 2.**
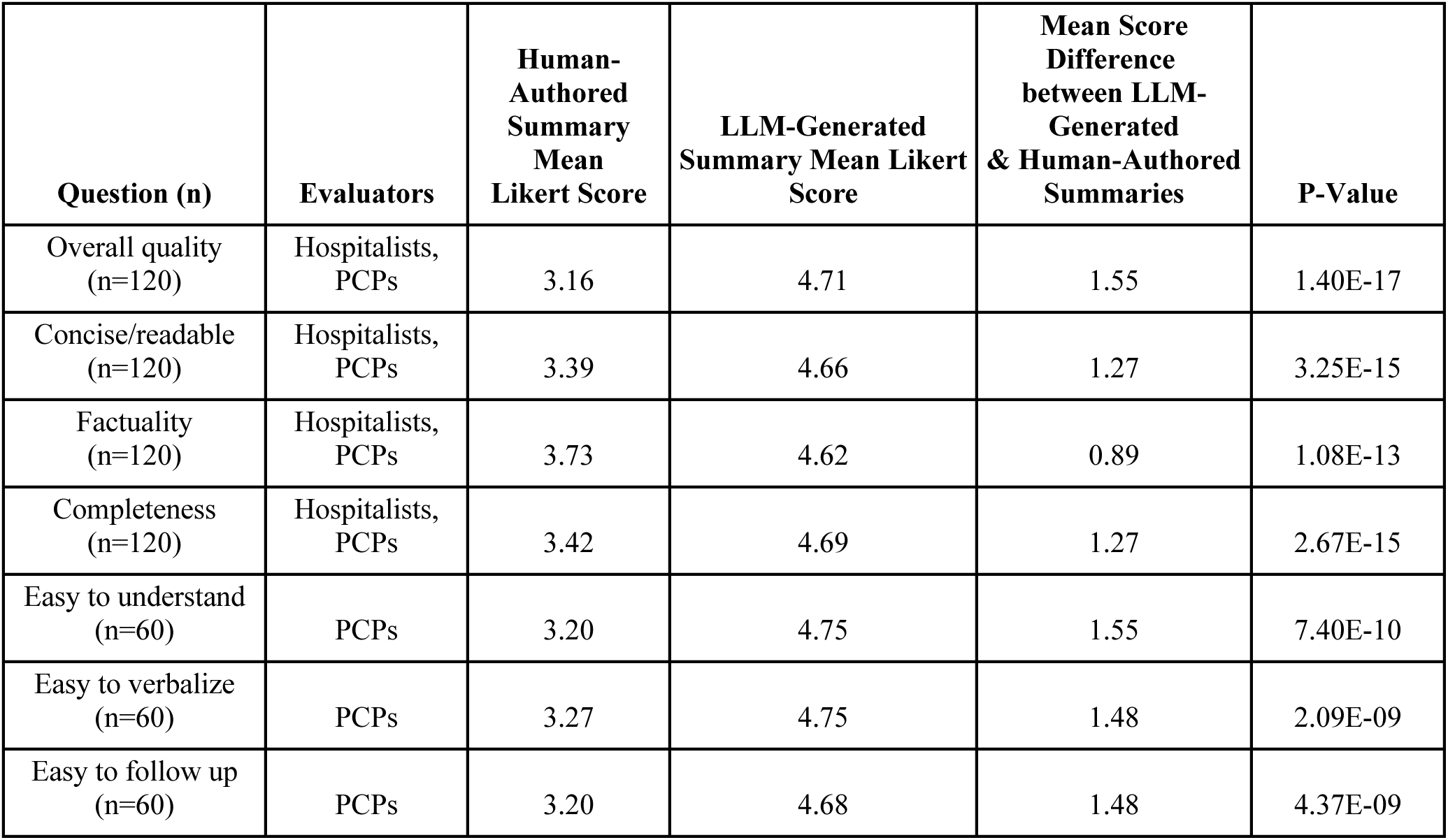
Mean survey results of physician reviewer ratings for human-authored vs. LLM-generated discharge summaries, based on a 1-5 Likert rating scale.

| Question (n) | Evaluators | Human-Authoried Summary Mean Likert Score | LLM-Generated Summary Mean Likert Score | Mean Score Difference between LLM-Generated & Human-Authoried Summaries | P-Value |
| --- | --- | --- | --- | --- | --- |
| Overall quality (n=120) | Hospitalists, PCPs | 3.16 | 4.71 | 1.55 | 1.40E-17 |
| Concise/readable (n=120) | Hospitalists, PCPs | 3.39 | 4.66 | 1.27 | 3.25E-15 |
| Factuality (n=120) | Hospitalists, PCPs | 3.73 | 4.62 | 0.89 | 1.08E-13 |
| Completeness (n=120) | Hospitalists, PCPs | 3.42 | 4.69 | 1.27 | 2.67E-15 |
| Easy to understand (n=60) | PCPs | 3.20 | 4.75 | 1.55 | 7.40E-10 |
| Easy to verbalize (n=60) | PCPs | 3.27 | 4.75 | 1.48 | 2.09E-09 |
| Easy to follow up (n=60) | PCPs | 3.20 | 4.68 | 1.48 | 4.37E-09 |

Physician reviewer ratings were more consistent for LLM-generated summaries than human-authored summaries. The mean absolute deviation in Likert scores for the LLM-generated summaries was less than half that of the human-authored summaries (0.49 vs 1.07, p = 0.001). Similarly, the exact match percentage, or proportion of paired survey responses where both reviewers assigned the same Likert score, for the LLM-generated summaries was more than double that of the human-authored summaries (57% vs 27%), and only 4% of the paired LLM-generated summary ratings had a difference of more than 1 point, versus 27% for the human-authored summaries.

Across 60 encounters containing 138 error annotations (47 inaccuracies and 91 omissions), there were fewer errors per LLM-generated summary than per human-authored summary (mean count [SD] 0.42 [0.98] vs 0.73 [1.34], p = 0.011). Mean potential and likelihood of harm score distributions between LLM-generated and human-authored summaries were similar (1.46 [1.84] vs 1.73 [1.89], p = 0.380 for harm potential; 2.30 [2.51] vs 2.11 [2.52], p = 0.648 for harm likelihood). Within error categories, there was a lower rate of omissions with LLM-generated vs. human-authored summaries, but no difference in the rate of inaccuracies (0.26 [0.72] vs. 0.5 [1.0], p = 0.004 for omissions; 0.16 [0.54] vs 0.23 [0.72], p = 0.45 for inaccuracies). Figure 1 shows the error counts and distributions of harm scores for both human-authored and LLM-generated discharge summaries.

**Figure 1.**
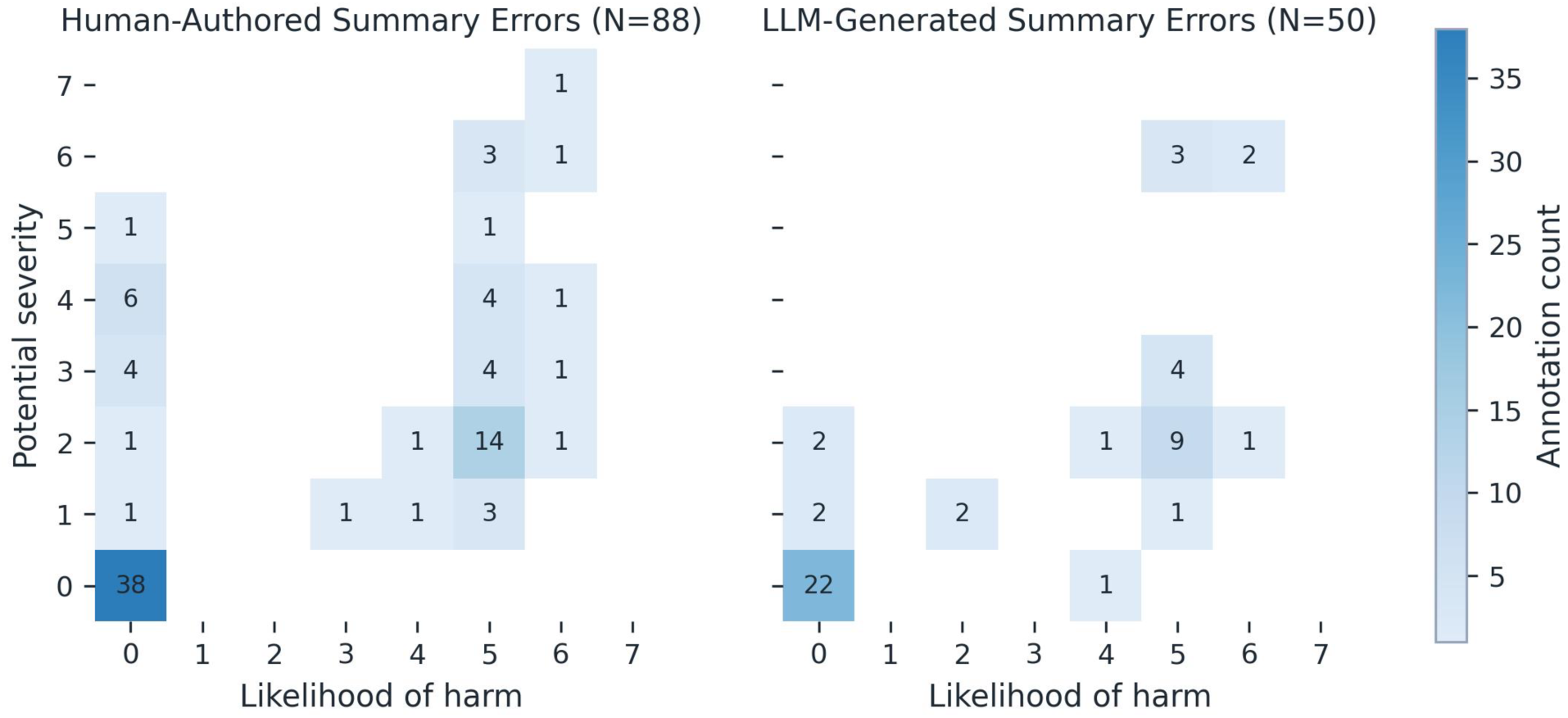
Heat map of physician reviewer annotated error counts and harm ratings by summary type. Each cell counts error annotations with the corresponding potential-severity and harm-likelihood ratings on the 0-7 scales.

For both human-authored and LLM-generated summaries, length of stay was not significantly correlated with mean Likert score for either summary type (spearman rho = 0.027, p = 0.837 for human-authored summaries; 0.138, p = 0.292 for LLM-generated summaries). Figure 2 shows the relationship between mean survey ratings and length of stay for both human-authored and LLM-generated discharge summaries.

**Figure 2.**
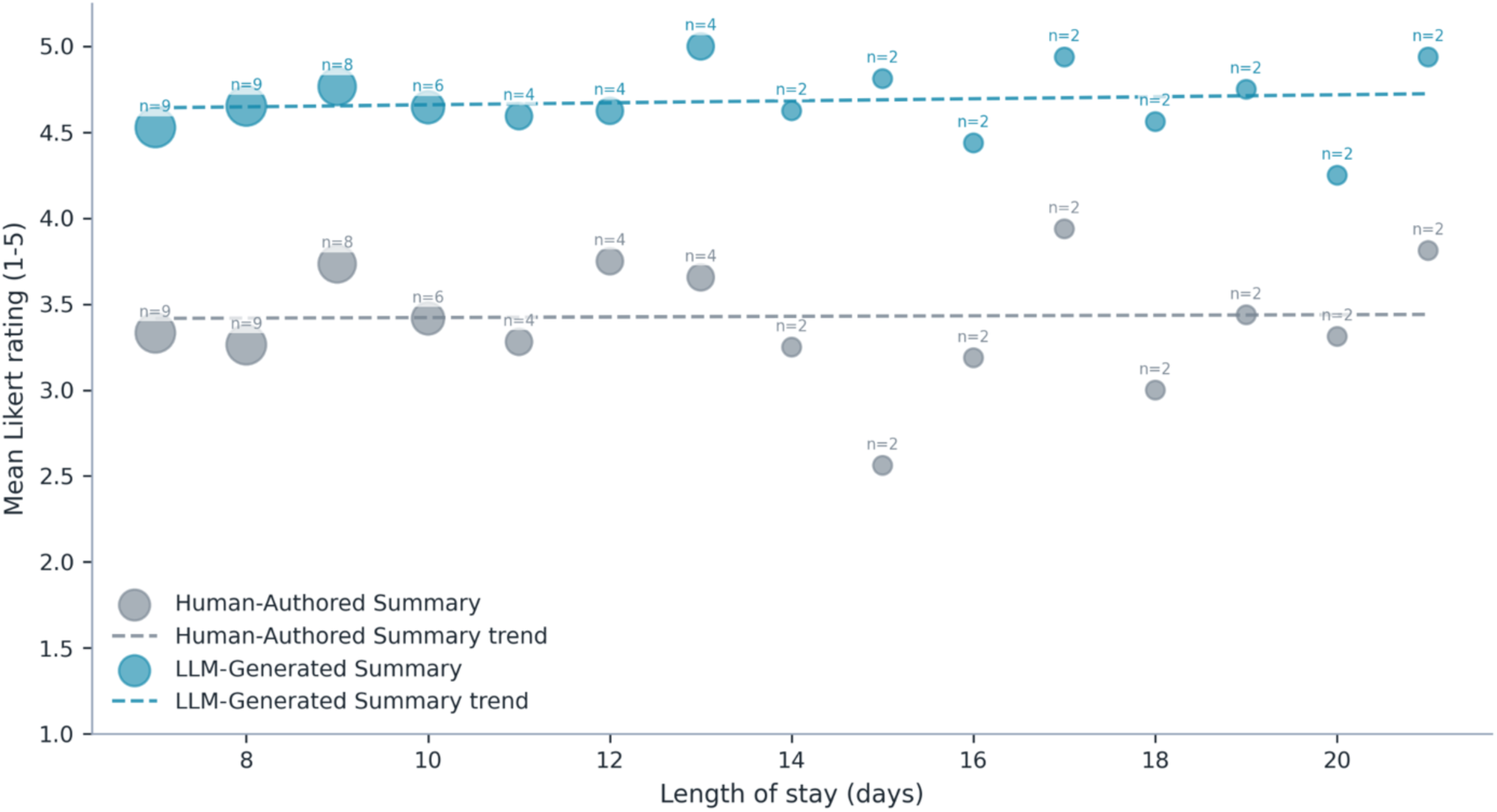
Scatterplot comparing mean survey ratings to length of stay for both human-authored and LLM-generated discharge summaries (n = 60). Individual values of n above each data point indicate the number of encounters available to calculate the average.

### Qualitative Themes

Themes identified from the debriefing sessions are shown in Supplementary Table S4. During the debriefing sessions, physician reviewers shared their insights comparing AI (LLM)-generated and human-authored summaries. Qualitative data concurred with the quantitative results. Specifically, PCP and hospitalists mentioned the following as main reasons underlying a preference for LLM-generated summaries: reduction of cognitive burden, higher consistency of summary qualities, and clearer presentation of critical information such as treatment rationale and unresolved follow-up issues. Most physicians found LLM-generated summaries lightened their cognitive burden in reading discharge summaries. One PCP noted that “AI summaries were just, obviously, I felt more understandable, more succinct, better organized. It told the story. It was easier to read rather than just kind of piecing through it and trying to understand.” Whereas for human-authored discharges, physicians sometimes “spend like an hour or two… scouring the chart, trying to figure out what happened… and can’t even figure out what happened”.

Physician reviewers noted consistency in the quality of LLM-generated summaries. One PCP mentioned that for human-authored summaries, they are “physician dependent” such that “some were better than others…some were quite detailed while others were just maybe three paragraphs. While for LLM-generated summaries, they were “more uniform…more understandable, more succinct, better organized.” Another hospitalist concurred with the PCP by noting that “[the main difference] is the inconsistency between physicians… [for human-authored summaries] there can be redundancy… there are so many things that are omitted that are important…Whereas the AI summaries are concise and there was no redundancy”.

Another reason for physician reviewers’ high ratings for LLM-generated summaries concerned the clarity of critical health information. Physician reviewers mentioned that treatment rationale was key information that they look for in the discharge summaries. One PCP mentioned an instance where the LLM-generated summary included treatment rationale whereas the corresponding human-generated summary lacked this information. Physicians also mentioned unresolved follow-up issues as key discharge summary information. One hospitalist noted that LLM-generated summaries did a better job presenting information regarding unsolved follow-up issues: “what I loved [about LLM-generated summaries], is that anything that was still pending was acknowledged and had a comment on who is appropriate to follow up with [for] these results…that is something that was essentially not found in any of the human-written summaries”.

### Provider-Specific Differences in Discharge Summary Evaluation

#### Quantitative Findings

The difference in scores between LLM-generated and human-authored summaries by provider type and survey question is shown in Figure 3. The mean score differences were higher for PCPs than hospitalists for readability/conciseness and completeness (mean [SD] 1.48 [1.28] vs 1.05 [1.14], p = 0.02 for readability/conciseness; 1.47 [1.38] vs 1.07 [0.97], p = 0.02 for completeness; Supplementary Table S6). Between providers, mean harm score distributions were similar for harm potential (mean [SD] 1.65 [1.83] vs 1.58 [2.02], p = 0.675), but PCPs assigned higher likelihood of harm than hospitalists (2.66 [2.51] vs 0.83 [1.98], p = 5.33e-4). PCPs and hospitalists identified similar counts of inaccuracies per summary (mean [SD] 0.40 [0.827] vs 0.38 [1.03], p = 0.664), but PCPs identified more omissions than hospitalists (1.3 [1.84] vs 0.22 [0.585], p = 1.16e-5). Overall, hospitalists and PCPs identified similar numbers of inaccuracies (49% vs. 51%), but PCPs identified most omissions (86% vs. 14%).

**Figure 3.**
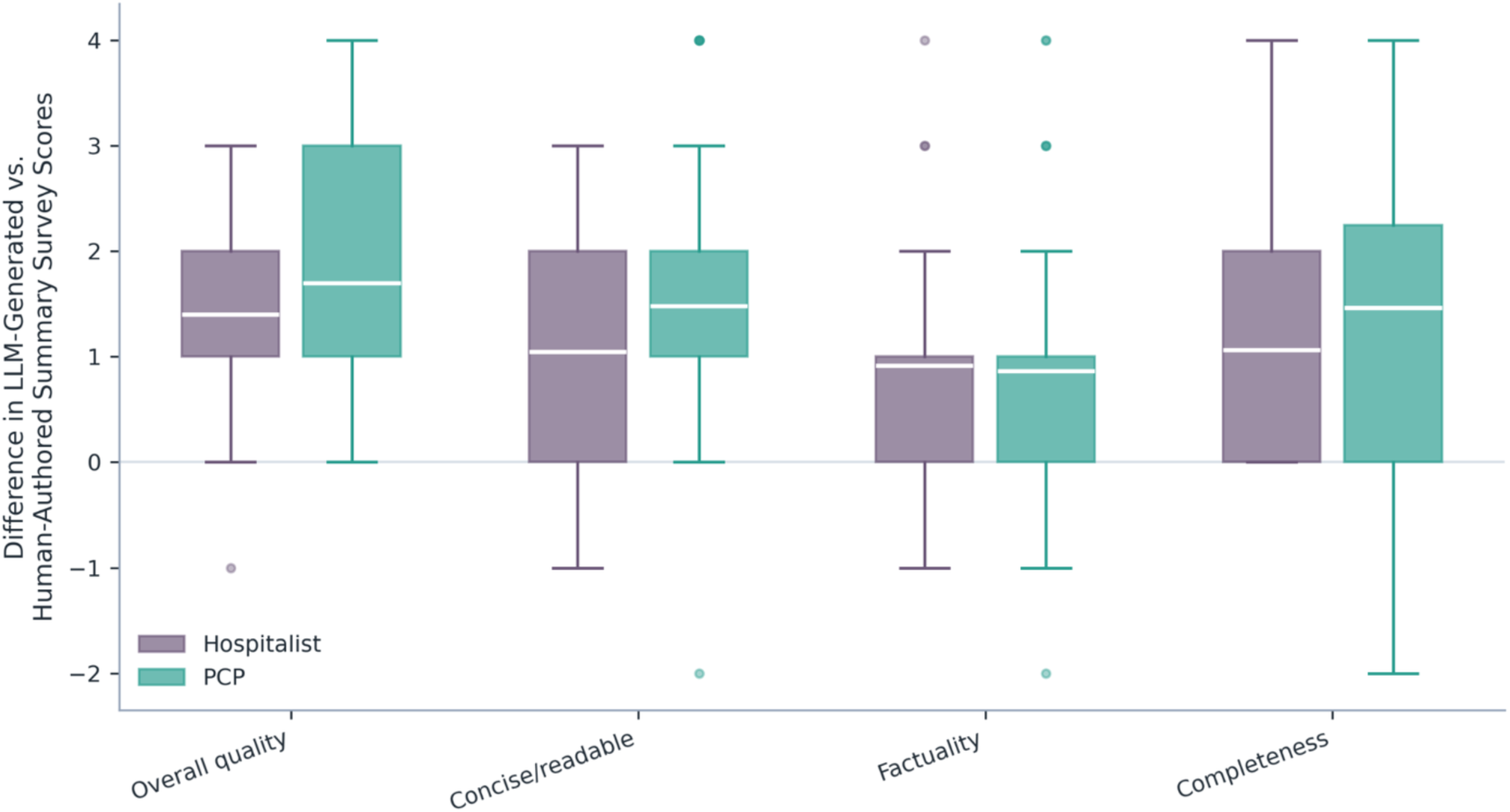
Difference in LLM-generated vs. human-authored survey scores by provider type and survey question (n = 120). Outliers are shown as individual points but excluded from mean, IQR and range calculations.

#### Qualitative Themes

To better understand the differences in ratings between PCPs and hospitalists, we probed physician reviewers’ own interpretations during the debriefing sessions. PCP reviewers clearly attributed higher harm likelihood scores to their long-term experience and view of patients. They explained that the reason why PCPs rated harm likelihood much higher was because they had experienced and had to deal with many medical complications after discharge. One PCP shared that “… because we’ve experienced, you know, somebody sent home on 2 anticoagulants…blood thinners…excessive psychiatric medications, too much diuretic. We see what happens. And often we’re the ones that have to deal with the complications.” There were also differences noted in risk tolerance between inpatient and outpatient care settings. One hospitalist noted that “(PCPs) throw the dice every day… So they probably have seen a lot more stuff than I’ve seen… So, like, they’ve seen a lot more of like bad stuff in non-controlled settings.”

Differences in provider workflows also emerged as an important theme. For PCPs, they described their workflow in reviewing discharge summaries for transition of care, and the need to request records and review details within individual EHR notes when discharge summaries were insufficient. One PCP shared that “I read the discharge summary, the medication reconciliation in there, and then I more often than not don’t trust it and go back and I look at vital signs, labs, micro and imaging from the hospitalization, go over it myself.” From the hospitalist perspective, the focus was on reviewing and synthesizing historically important information from the EHR. As a hospitalist noted, “I know I took over a patient once that had been in the hospital for over 3 months without a good hospital course. And it took me two hours to go through that chart, just generate a discharge summary.”

### Evaluation of Incidental Finding Identification

#### Quantitative Findings

There were 31 LLM-identified incidental findings from radiology reports. Physician reviewers rated 93.5% of the findings as factually correct and 87.1% appropriate to be reported as an incidental finding on the discharge summary (Supplementary Table S7).

#### Qualitative Themes

Some notable differences in perspectives on incidental finding reporting emerged during debriefing sessions among PCPs and hospitalists. Several hospitalist reviewers expressed their favor for a broader inclusion of incidental findings. One noted that “As a hospitalist, I am covering my own bases, right? So I’m calling out everything (that could be an incidental finding) and then I let (the PCP) decide which ones he wants to follow or not.”

## Discussion

### LLM-Generated Summaries Improve Quality and Reduce Cognitive Burden

In this study of patients with hospitalizations between 7-21 days, our results suggest that LLM-generated discharge summaries outperformed human-authored summaries in producing more understandable, consistent, and actionable handoffs without introducing substantial errors. Across evaluations by both hospitalists and PCPs, LLM-generated summaries received higher ratings for overall quality, readability/conciseness, factuality, and completeness, and physicians indicated a preference for the LLM-generated summary in 95% of encounters. Importantly, PCPs, the intended recipients of discharge summaries for transition-of-care management, rated LLM-generated summaries higher for ease of understanding and verbalizing the major aspects of the hospital course to patients, and for supporting follow-up care. The narrower distribution of LLM-generated summary ratings is also notable, as it suggests that LLM-generated summaries may reduce the author-dependent variability that commonly affects human-authored discharge documentation. Feedback from the debriefing sessions suggest that these differences were meaningful because LLM-generated summaries reduced the cognitive burden of reconstructing chronological events, improved the consistency in quality across discharge summaries, highlighted rationale for treatment changes, and improved identification of unresolved issues requiring follow-up.

### Safety and Error Characteristics of LLM-Generated Summaries

Analysis of physician-annotated errors demonstrated that human-authored summaries contained more errors than LLM-generated summaries, with omissions representing the most common error type in both groups. Errors in human-authored summaries more often involved omission of clinically important details related to hospital management and transition-of-care planning, including treatments, medication discontinuation plans, relevant consultations and follow-up recommendations. In contrast, errors in LLM-generated summaries were generally narrower in scope and more commonly involved isolated omissions of medication details, follow-up timing, or contextual information such as relevant comorbidities prior to hospitalization. Importantly, there was no difference in the potential severity or likelihood for harm between errors in LLM-generated compared to human-authored summaries. These findings suggest that while LLM-generated summaries are not error-free and still require physician oversight, they do not produce more clinically meaningful errors than those that commonly occur in human-authored discharge summaries.

### LLM Performance for Long and Clinically Complex Hospitalizations

The stability of findings across 7 to 21-day hospitalizations have important implications, with LLM-generated summaries maintaining consistently superior performance despite increasing information volume. Longer encounters often produce larger quantities of notes, repetitive information, and complex clinical trajectories that are difficult for both discharging and receiving physicians to synthesize. Debriefing sessions suggest that these differences may translate into meaningful time savings and reduced cognitive burden in reconstructing the hospital course from fragmented documentation. Participants noted that with longer hospitalizations, time constraints often force clinicians to skim lengthy records, increasing the risk that important issues such as rationale for management decisions or unresolved follow up issues may be omitted. In addition, the methods used in this study demonstrate that LLMs can generate coherent discharge summaries even for EHRs exceeding the model’s context window, an important technical consideration given the large volume of documentation associated with longer hospitalizations.

### Differences Between PCP and Hospitalist Perspectives

The advantages of LLM-generated summaries appeared to be more pronounced among PCPs than hospitalists. Across both overall and paired evaluations, PCPs demonstrated larger differences in ratings favoring LLM-generated over human-authored summaries, particularly for overall quality and conciseness/readability, with less variability between PCP and hospitalist ratings for the LLM-generated summaries. PCPs also identified more errors, specifically omissions, in both human-authored and LLM-generated summaries than hospitalists, with higher scores for likelihood of harm. Debriefing sessions suggested several reasons for this difference. PCPs often perceived greater downstream harm potential from omissions or inaccuracies because they are responsible for longitudinal follow-up and management of post-discharge complications. As one PCP noted, “we see what happens, and often we’re the ones that have to deal with the complications,” contributing to higher perceived likelihood of medication or follow-up errors from incomplete discharge documentation. Additionally, workflow differences likely contributed, as PCPs described frequently needing to review inpatient notes to reconstruct the hospitalization when key details were missing from discharge summaries, while hospitalists described the challenge of condensing prolonged and fragmented hospitalizations into a concise narrative. PCPs may perceive greater value from LLM-generated summaries because their workflow requires directly acting on discharge documentation to provide ongoing care, whereas hospitalists are primarily focused on creating a summary for handoff. The prompt engineering in this study was designed with PCPs as the target audience, suggesting that incorporating PCP informational priorities and workflow needs into development may improve discharge summaries for the receiving clinicians who provide follow-up care.

### Incidental Finding Identification and Reporting

Our results suggest that LLMs performed well as a tool for identifying and summarizing incidental findings. Most incidental findings identified in LLM-generated summaries were judged to be both appropriate and correctly reported, and notably, reviewers did not annotate omission errors related to incidental findings from radiology studies. Debriefing sessions highlighted differences between hospitalist and PCP perspectives on incidental findings, with hospitalists favoring broader reporting to ensure abnormalities were documented and questioning whether LLMs might underreport incidental findings, while PCPs preferred selective reporting of actionable findings to avoid information overload. Importantly, the prompt for incidental findings was designed for specificity, where the LLM was directed to identify findings that were specifically flagged by radiologists for follow-up. However, both groups agreed that LLM-generated incidental finding summaries were beneficial for reducing the likelihood that clinically important findings requiring follow-up would be missed.

### Implications for Clinical Adoption and Oversight

Although both quantitative evaluation and qualitative debriefing indicated the tremendous potential of LLM-generated summaries, physicians cautioned against blind reliance on LLMs. When probed with final thoughts on adopting LLM-generated summaries in clinical workflows, physicians emphasized that LLM summaries should only function within a clinician-supervised context. Physicians in all three debriefing sessions strongly agreed that, as reflected in one hospitalist’s quote, “no matter what information is pre-populated or anything, it’s still the responsibility of the physician to make sure it’s correct.” Physicians specifically highlighted the importance of deliberate verification workflows for high-risk content such as medication changes, follow-up recommendations, and incidental findings. Moreover, some had cautionary comments on potential threats for widespread adoption of LLM-generated summaries in clinical workflows. One hospitalist cautioned that “I guarantee you no one’s going back and fact checking or reading everything to make sure that it’s fine… Because usually when something’s handed to you, you’re just going to take it as is and you give up your agency to kind of do some of the things that doctors do… I’m afraid that, you know, they’re not going to do any QA [quality assurance] to make sure that everything is correct."

## Limitations

This study has several limitations. First, it was conducted at a single academic medical center using one EHR environment and local documentation practices, which may limit generalizability to other health systems, specialties, or workflows. Second, the study evaluated physician perceptions of discharge summary quality and safety rather than downstream patient-centered outcomes such as timely completion of discharge summaries, readmissions, medication errors, or adverse events after discharge. Although clinician assessment is highly relevant for transition-of-care workflows, future prospective studies are needed to determine whether these perceived advantages translate into measurable improvements in patient outcomes and operational efficiency. Lastly, while qualitative debriefing sessions provided important contextual insights into workflow, trust, and clinician perceptions, we did not use formal qualitative research methodologies such as structured thematic coding or saturation analysis, and these findings should therefore be interpreted as exploratory.

## Conclusion

In conclusion, LLM-generated discharge summaries for longer hospitalizations were consistently rated higher than human-authored summaries across multiple performance metrics by both hospitalists and PCPs. The perceived benefits and quality advantages were particularly pronounced among PCPs, suggesting that designing discharge summaries around the informational priorities and workflow needs of receiving providers may meaningfully improve transition-of-care communication for those who provide post-hospitalization care. These findings support the potential for thoughtfully implemented, LLM-assisted summarization to reduce cognitive burden and better support transitions of care, while maintaining the need for clinician oversight and verification.

## Acknowledgments

We would like to take an opportunity to acknowledge some additional people who were instrumental in this study.

First, we would like to thank Fang Wang, a Clinical Research Data Analyst/Architect with the Research Computing & Informatics (RCI) team at Stony Brook University, for her efforts in extracting and the raw EHR dataset used in this study.

Second, we would like to thank Erich Bremer, MSc, the Director of Applied Informatics, and Joe Balsamo, a Supervising Programmer Analyst, from the Stony Brook Medicine Department of Biomedical Informatics, for their efforts in setting up the servers and access necessary for us to develop the HIPAA-compliant full-stack web application to collect review data from the clinicians.

This work was a true team effort; we could not have done it without everyone’s enthusiastic support. Thank you.

