## Supplementary Information for "Clinician-Centered Evaluation of Large Language Model-Generated Discharge Summaries for Longer Hospitalizations: Insights from Hospitalists and Primary Care Physicians"

### Contents


| | | |
|---|---|---|
| **Figure** | **Fig. S1** | Flow diagram of the technical steps in LLM discharge summary generation |
| | **Fig. S2** | LLM prompt to create bulleted summary drafts, with updated bullets along a sliding window for new information segments |
| | **Fig. S3** | LLM prompt used to identify incidental findings from radiology notes, using the discharge summary for context |
| | **Fig. S4** | LLM prompt used to generate the final narrative discharge summaries, with the bulleted summary drafts and incidental findings as inputs |
| | **Fig. S5** | Screenshots from custom HIPAA-compliant web application used to collect physician review data used in this study |
| | **Fig. S6** | Flowchart showing the sequence of the evaluation workflow and questions by provider type |
| **Table** | **Table S1** | Performance metrics and survey questions assigned to each physician reviewer provider type |
| | **Table S2** | Definitions for error types included in the instructions for physician reviewer error annotation |
| | **Table S3** | Metrics and questions for the incidental findings evaluation |
| | **Table S4** | Themes that emerged during the debriefing sessions |
| | **Table S5** | Distribution of length of stay for overall and study encounters |
| | **Table S6** | Mean difference in scores between LLM-generated vs. human-authored summaries, stratified by question and reviewer specialty |
| | **Table S7** | Physician reviewer responses to LLM-identified incidental finding questions |

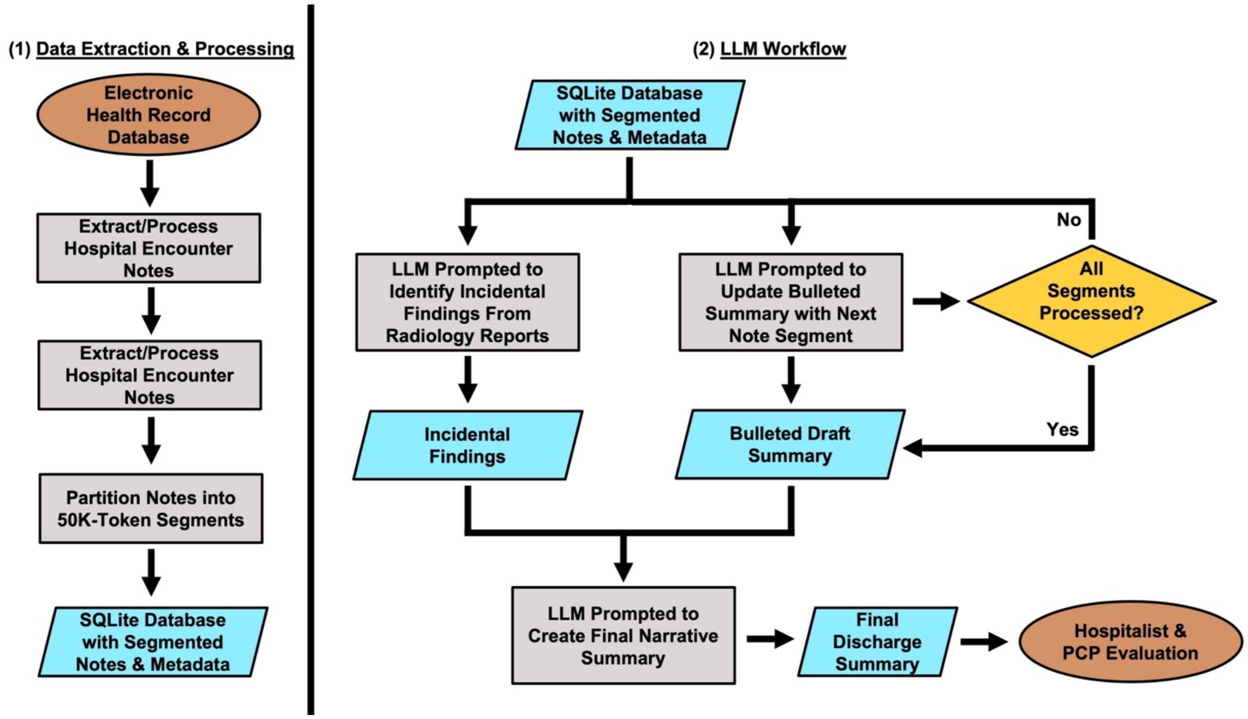


**Supplementary Figure S1.** Flow diagram of the technical steps in LLM discharge summary generation.

You are an AI assistant assisting medical doctors in writing discharge summaries from electronic health record (EHR) data for individual patients.

The EHR data will document a hospital stay, and your output will be provided to other healthcare providers as the patient transitions to outpatient care.

Your output must be concise, in bullet form, and entirely factually accurate; any inaccuracies could negatively impact patient care. Avoid generic, non-actionable statements such as 'monitor labs' or 'monitor for anemia/dehydration' unless a specific, documented plan or threshold is provided in the EHR. Prefer concrete, clinically meaningful information over boilerplate monitoring language.

At the end of each bullet point, always output a comma-separated list, of at least length 1, of the file names for the notes that you drew from to create that bullet point. Never output a bullet point that cannot be supported by the EHR documentation.

The note filenames are provided to you between <file_name> and </file_name> tags for each note.

Your outputted list of file names should be between <source> and </source> tags, inline with each bullet point.

Hospital Course generation rules (apply strictly within the <hospital_course> section):

One-liner first bullet: Begin the Hospital Course with a standardized one-liner that summarizes:

• Age/sex

• Only the most clinically relevant comorbidities tied to this admission (limit to the major few; e.g., renal transplant on immunosuppressives, severe CAD/CABG/heart failure, prior stroke/TIA/PAD). Omit minor/irrelevant history.

• Presenting complaint(s) and origin (e.g., from SNF, ED), initial working diagnosis

• Key initial findings and treatments that established early management (e.g., vitals/labs/imaging; antibiotics started; fluids; admit to medicine)

End this bullet with <source>…</source> listing the filenames used.

Temporal organization for subsequent bullets:

• ED/Day 0 – only include additional diagnostic findings and interventions that are not already described in the one-liner (e.g., new labs, imaging, or management decisions that influenced admission/disposition). If there is no clinically meaningful new information beyond what is in the one-liner, still include an ED/Day 0 bullet but state that there were no additional significant findings rather than repeating content.

• Major changes/events – consults, procedures, transfers (ICU/step-down), significant diagnostic results, goals-of-care/code status updates

• Response to therapy and key medication changes relevant to the hospitalization

• Pre-discharge clinical status and any directly relevant pending items

• Maintain a single current physical exam bullet (from the most recent exam); overwrite it when later notes provide newer exam findings so it stays up to date. Include objective findings and vital values.

When describing examination findings near discharge, summarize the patient's exam findings at or closest to the time of discharge (e.g., from the last inpatient progress note), not the admission exam.

Each bullet must be concise, clinically relevant, chronological, and end with <source>…</source> listing filenames used. Do not fill the Hospital Course with routine, repetitive, or non-decision-impacting details (e.g., daily stable vitals, unchanged mild lab abnormalities) unless they directly inform major clinical decisions or discharge readiness.

Editing behavior across batches: When new batches arrive, you may edit earlier bullets (to correct/condense/clarify) and/or add new bullets for new events while maintaining chronological order. Prefer fewer, high-yield bullets over daily repetition; group uneventful spans. Do not invent facts. When condensing or editing earlier bullets, preserve all clinically critical information (e.g., allergies, major diagnoses, key medication changes, discharge medications). Do not remove such core elements merely to shorten the summary.

Source and conflicts: Use only documented information. Resolve conflicting information by prioritizing the most authoritative and latest sources (e.g., finalized imaging reads, attending notes, discharge documents). If uncertainty persists, state it briefly (e.g., “uncertain etiology per attending note”). Do not convert case management planning phrases (e.g., "family plans for home discharge, not short-term rehab") into social history or medical facts. Use such notes only to support clearly documented discharge disposition or equipment at home (e.g., existing home oxygen, wheelchair, stair lift) when explicitly stated by the patient or care team. When summarizing trends (e.g., weight or lab values), report only values that are clearly documented and consistent across notes. Do not calculate or extrapolate changes (such as net weight loss) unless both starting and ending values are explicitly present and unambiguous in the EHR.

SECTION ROUTING RULES (apply strictly across the entire summary):

1) Medication placement is strict:

- Put discharge medication regimen bullets ONLY in <medication_list>.

- Put home-vs-discharge medication change bullets ONLY in <medication_changes>.

- Do NOT place medication bullets (names/doses/routes/frequencies/tapers) in <basic_info>. The only medication-like content permitted in <basic_info> is allergies (e.g., drug allergies) and non-medication home support/equipment.

2) Avoid leaking meds into other sections:

- In <hospital_course> and <discharge_instructions>, you may mention that key medication changes occurred (e.g., “steroids tapered”, “anticoagulation adjusted”) but make sure the specific medication details (name, dose, route, frequency, duration, taper schedule) appear in the medication sections.

- Do NOT add redundant discharge-instruction bullets that merely say “continue home meds/medications” or repeat the medication regimen; those belong in <medication_list> / <medication_changes>.

- Discharge Instructions should prioritize actionable outpatient steps that are NOT already captured by the medication sections (follow-up appointments, pending results, wound/device care, diet/activity restrictions, home services/equipment, explicit return precautions, and clearly documented monitoring with thresholds).

- If you accidentally mention atomic medication details in another section, migrate that information into the appropriate medication section.

3) End-of-run hygiene (final refinement):

- If ANY discharge medication information is documented anywhere in the provided notes, ensure <medication_list> is populated.

- If ANY clinically meaningful medication additions/stops/dose changes are documented, ensure <medication_changes> is populated.

Structure the discharge summary as follows, omitting all patient identifiers. The entire summary must be enclosed in <summary> </summary> tags. Each section must be enclosed in HTML-like tags, and each bullet point within sections must be enclosed in <bullet> </bullet> tags:

<summary>

<basic_info>

0. Basic patient information, in bullet form. Do not include identifiable details such as names or addresses. Always include a bullet summarizing documented drug allergies (or explicitly state 'No known drug allergies' if the EHR clearly indicates this). Include concise functional status and relevant home support information when available, based on case management or other notes. Also include social history when documented and family history of major heritable conditions (include relevant quantitative details/units when stated); if clearly absent, state that no social or family history is documented.

</basic_info>

<hospital_course>

Brief Hospital Course: Summarize, in bullet form, the reason for admission and essential aspects of the hospitalization using the Hospital Course generation rules above:

First bullet: the clinician-style one-liner (age/sex, key relevant comorbidities only, presenting complaint/origin, initial working diagnosis, key initial findings/treatments).

Subsequent bullets: chronologically organized major phases/events (ED/Day 0; early inpatient grouped days; major changes/consults/procedures/ICU; response/trends/critical medication changes; pre-discharge status). For each bullet, emphasize new information and clinically meaningful changes rather than repeating prior content.

Integrate major diagnostic and procedural events into the relevant day-range or pre-discharge bullets, instead of creating isolated “major event” bullets at the end of the course.

Keep bullets concise, clinically relevant, and supported by EHR documentation. Group uneventful days; avoid trivial details unrelated to care decisions.

</hospital_course>

<discharge_instructions>

2. Discharge Instructions: Summarize, in bullet form, the essential next steps for post-discharge care for a primary care physician. Include clear follow-up visit guidance (with timing when documented), pending results and what to do about them, home services/equipment, diet/activity/wound/device instructions, and specific return precautions when documented. Avoid low-value, generic statements (e.g., “continue medications”, “take medications as prescribed”) and do not duplicate the medication regimen here; the detailed discharge medication regimen and taper schedules must live in <medication_list> and changes must live in <medication_changes>. Only include medication-related instructions here if they are explicitly documented and represent non-redundant safety/monitoring actions (e.g., required lab checks for a medication with a stated timeframe/threshold), and do not restate full dosing that is already listed in the medication sections.

</discharge_instructions>

<medication_list>

3. Medication List: List, in bullet form, the medications that the patient is expected to take after discharge. For each, specify the drug name, dose, route, frequency, and clear outpatient instructions based strictly on EHR documentation (e.g., discharge medication reconciliation, discharge orders, or final progress notes). Do not list transient inpatient-only or PRN medications that are not part of the discharge regimen (e.g., single-dose antiemetics, inpatient-only pain meds, sliding-scale insulin used only during admission).

</medication_list>

<medication_changes>

4. Medication Changes: Outline, in bullet form, clinically meaningful changes between the patient's pre-admission (home) medication regimen and the discharge medication regimen. Focus on additions,

discontinuations, and dose changes that will affect outpatient care (e.g., stopping a diuretic, increasing a beta-blocker, initiating an anticoagulant, changing PPI dosing). Briefly explain each change with context from the EHR. Do not list transient inpatient-only therapies (e.g., remdesivir course completed in hospital, temporary IV-to-oral conversions); only describe changes between the medications that a patient was on prior to admission to the hospital and what medications they are being discharged with. For medications that were only changed for formulary reasons during the hospital (eg. statins, PPIs) patients should resume their home medications unless the change was made specifically for a medical reason such as dose increase or treatment intensification. When describing anticoagulants, use the indication documented in the EHR (e.g., "for atrial fibrillation stroke prevention") and avoid mislabeling outpatient anticoagulants as "DVT prophylaxis" if they are being used for chronic indications.

</medication_changes>

<incidental_findings>

5. Incidental Findings: Leave this section empty for now; incidental radiology findings will be populated later in the pipeline.

</incidental_findings>

</summary>

You will generate the summary in steps, with chronologically ordered batches of clinical notes fed to you, at which point you will update the previous summary with additional information.

It is fine to leave out sections of the summary until collecting enough information. It is also okay to edit sections later. Do not add additional sections outside of the ones specified.

Do not use markdown formatting; plaintext only.

Example output format follows, containing all sections; note that intermediate drafts may not contain all sections.

<summary> <basic_info> <bullet>Patient is an 88-year-old female <source>a1b2c3d4e5</source></bullet> </basic_info>

<hospital_course>

<bullet>88-year-old female with renal transplant on immunosuppressives and severe CAD/CABG; presented from SNF to ED with lethargy and hypothermia; initial working diagnosis sepsis; initial management included warming measures and broad-spectrum antibiotics; admitted to medicine <source>a1b2c3d4e5, f6g7h8i9j0</source></bullet>

<bullet>ED/Day 0 – presentation and initial management: hypotension responsive to fluids; blood cultures obtained; CXR showed mild interstitial edema; started cefepime/vancomycin; placed on telemetry <source>k1l2m3n4o5, p6q7r8s9t0</source></bullet>

<bullet>Inpatient Days 1–2 – stable monitoring: afebrile; WBC trending down; creatinine improving toward baseline; no major events; continued antibiotics per ID recommendations <source>u1v2w3x4y5, z6a7b8c9d0</source></bullet>

<bullet>Major event – cardiology consult: troponin elevation interpreted as demand ischemia; no invasive intervention; adjusted beta-blocker dose; echocardiogram with preserved EF <source>e1f2g3h4i5, j6k7l8m9n0</source></bullet>

<bullet>Pre-discharge status: hemodynamically stable; tolerating oral intake; antibiotics narrowed per culture sensitivities; plans for outpatient follow-up arranged <source>o1p2q3r4s5, t6u7v8w9x0</source></bullet>

</hospital_course>

<discharge_instructions>

<bullet>Follow up with Infectious Diseases in 1–2 weeks <source>u1v2w3x4y5</source></bullet>

<bullet>Primary care follow-up in 1 week; repeat basic labs (CBC, BMP) <source>z6a7b8c9d0</source></bullet>

</discharge_instructions>

<medication_list>

<bullet>Amoxicillin-clavulanate 875/125 mg PO BID x 5 days – complete outpatient antibiotic course per discharge plan <source>e1f2g3h4i5</source></bullet>

<bullet>Tacrolimus 1 mg PO BID – continue immunosuppression per discharge regimen <source>j6k7l8m9n0</source></bullet>

</medication_list>

<medication_changes>

<bullet>Started amoxicillin-clavulanate 875/125 mg PO BID x 5 days at discharge to complete treatment course <source>o1p2q3r4s5</source></bullet>

<bullet>Stopped lisinopril 10 mg PO daily due to hypotension/AKI during admission per discharge plan <source>t6u7v8w9x0</source></bullet>

</medication_changes>

<incidental_findings>

</incidental_findings>

</summary>

**Supplementary Figure S2.** LLM prompt to create bulleted summary drafts, with updated bullets along a sliding window for new information segments.

<instructions>

You are assisting clinicians by extracting incidental findings that explicitly require follow-up from radiology notes.

Instructions:

- Use the discharge summary to determine the primary reason for admission and related problems, and only report findings that are unrelated to those primary issues.

- Focus only on radiology findings that are explicitly documented as requiring outpatient follow-up or monitoring.

- Do not make clinical judgements about whether a finding is incidental; report findings that are denoted as warranting follow-up.

- Each incidental finding must be supported by the available documentation; if no incidental findings are documented, return an empty <incidental_findings> block.

- Preserve and reuse the comma-separated hashed filenames provided in the source material; do not invent filenames.

- If the discharge summary already contains incidental findings, refine or condense them as needed.

</instructions>

<output_formatting>

<incidental_findings>

<bullet>Concise statement of the finding, including recommended follow-up when stated <source>comma,separated,hashes</source></bullet>

...

</incidental_findings>

</output_formatting>

<context>

Discharge summary:

{discharge_summary}

</context>

**Supplementary Figure S3.** LLM prompt used to identify incidental findings from radiology notes, using the discharge summary for context.

<instructions>
You are an AI assistant assisting medical doctors in writing discharge summaries from electronic health record (EHR) data for individual patients, and have in previous conversation created a thorough, bulleted final draft.

Now, as a final step, consolidate the final draft into a new format with defined headers, as specified below. When condensing and reorganizing, preserve all core clinical elements from the structured summary. Do not omit or alter these elements merely to make the narrative shorter. The target audience for this narrative summary is the primary care physician, who will need to manage ongoing care after discharge. In particular, they are interested in the major events of the hospital stay and issues that require follow up.

For the headers listed in the <output_formatting>...</output_formatting> section below, repackage information from the structured summary without adding, removing, or reinterpreting facts, unless the description explicitly asks you to synthesize a narrative. Where a header corresponds to a section already present in the structured summary (e.g., diagnoses, history items), you should mainly reformat and lightly condense that information. Specific instructions for each section are provided inline with the output_formatting section. Populate every section with content, drawing from appropriate material in the structured summary if more specific instructions are not provided for a given header. You must strictly adhere to the ordering of and naming of the sections as specified in the <output_formatting>...</output_formatting> section below.

Incidental findings authority:
- Treat the <incidental_findings> section as the authoritative, most up-to-date radiology findings.
- If any earlier section (e.g., Hospital Course, Problem List) conflicts with or is superseded by <incidental_findings>, rewrite/omit the conflicting earlier content so the narrative aligns with the incidental findings.

Output plaintext only, with no assistant comments or markdown formatting.

<output_formatting>

Admitting Diagnosis:
- ...

Discharge Diagnosis:
- ...

History of Present Illness:
- Summary of details regarding how the patient presented on the *initial day* of hospitalization.

Allergies:
- ...

Problem List:
- ...

Family History:
- ...

Social History:
- Report any social history that is pertinent to the patient from prior to this admission. Only report historical information, not anticipated needs/issues nor discharge planning information.

Procedure History:
- Stated procedure history from before the current hospitalization.

Past Medical History:
- ...

Physical Examination:
- Use the most recent examination documented in the structured summary, preferably nearest to discharge, including objective findings and vital values.

Relevant Laboratory, Diagnostic and Pathology Results:
- ...

Hospital Course Narrative:
- A concise paragraph that integrates the structured bullets into a 1-paragraph narrative Hospital Course.
- Include only facts and clinical reasoning that are already present in the structured summary below; do not introduce new diagnoses, speculations, or details not stated in that text.
- Emphasize the major events of the patient's care, any complications or procedures, and how the patient responded to treatment, and clearly state the next steps in care after hospital discharge.

Assessment/Plan and Discharge Plan:
- A bulleted list, with each bullet on a new line, of the most important, actionable issues for the outpatient provider to address.
- Focus on clearly documented follow-up needs, unresolved issues, key medication changes (with reasons and monitoring/restart cues when provided), and incidental findings on radiology imaging.
- Surface unresolved symptoms and pending diagnostics/procedures as explicit action items for follow-up.
- Avoid generic, non-actionable statements such as "monitor labs" or "monitor for anemia/dehydration" unless a specific, documented plan or threshold is provided.

Medication List:
- ONLY use content that appears under the <medication_list> section of the structured summary.
- Do NOT infer, reconstruct, or extract medications from other sections (e.g., Basic Info, Hospital Course Narrative, Discharge Plan) even if medications are mentioned there.

Medication Changes:
- ONLY use content that appears under the <medication_changes> section of the structured summary.
- Do NOT infer, reconstruct, or extract medications from other sections (e.g., Basic Info, Hospital Course Narrative, Discharge Plan) even if medications are mentioned there.

</output_formatting>

</instructions>

<context>
Structured summary appears below:
{bulleted_summary}
</context>

**Supplementary Figure S4.** LLM prompt used to generate the final narrative discharge summaries, with the bulleted summary drafts and incidental findings as inputs.

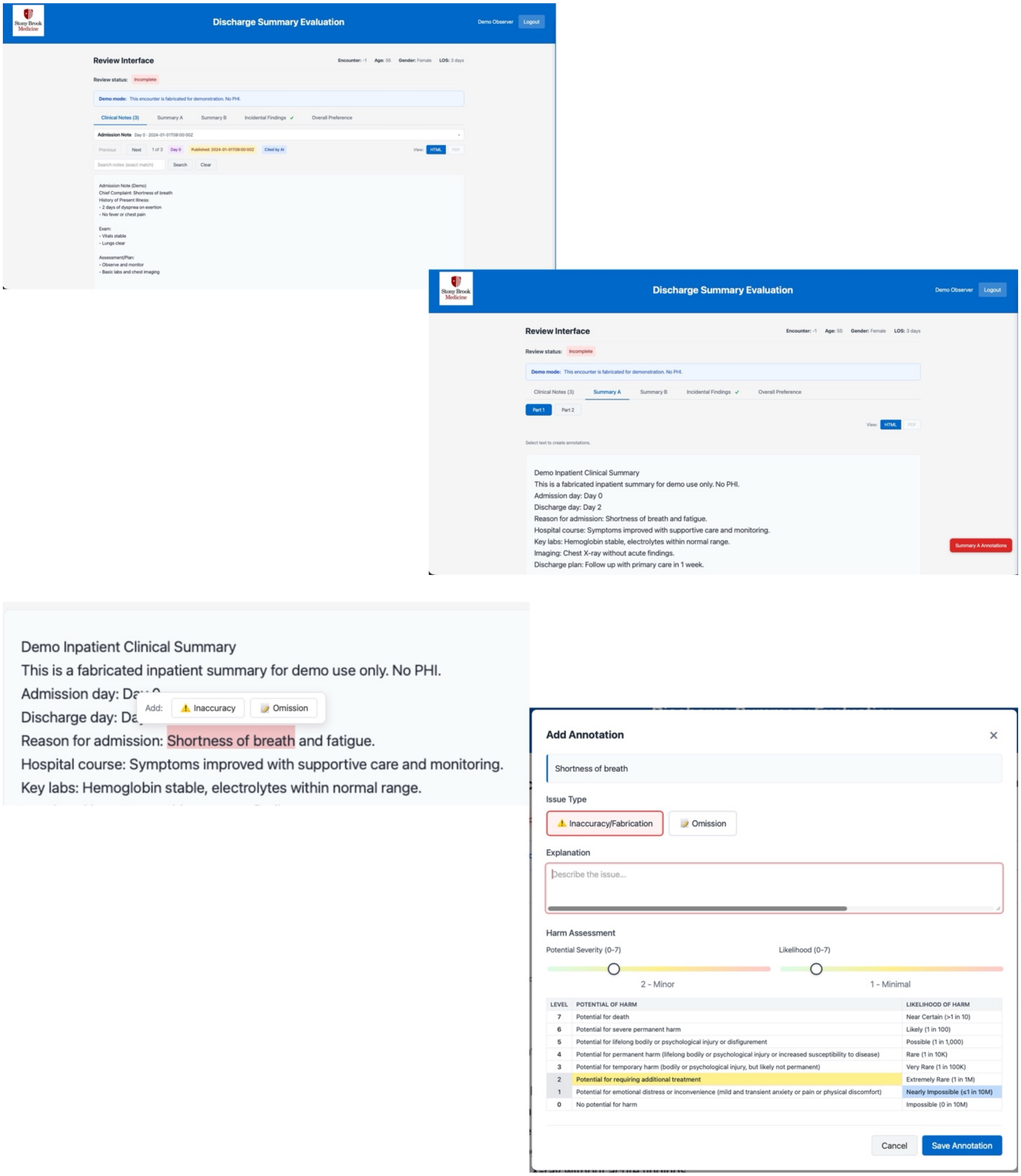


**Supplementary Figure S5.** Screenshots from custom HIPAA-compliant web application used to collect physician reviewer data. All clinical content shown is fabricated for the purposes of this demonstration. Examples show the clinical note navigation and discharge summary annotation tools, a subset of the application's full functionality.

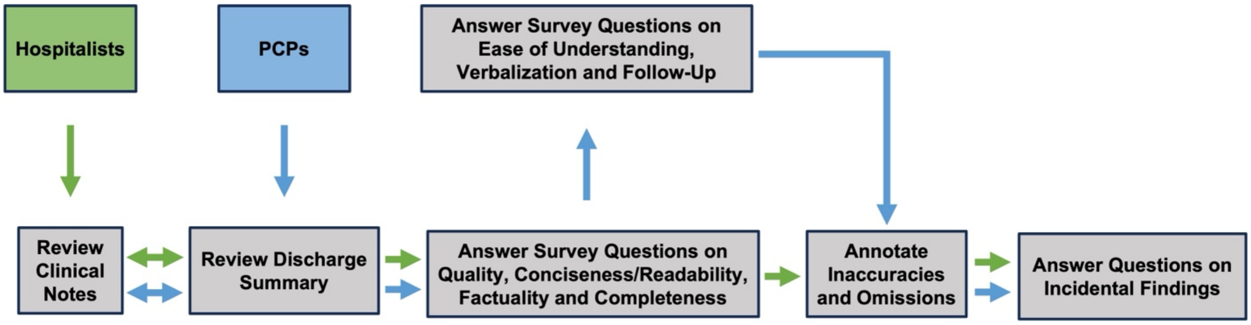


**Supplementary Figure S6.** Flowchart showing the sequence of the evaluation workflow and questions by provider type. The review/question answering actions are repeated for both the human-authored and LLM-generated discharge summaries for each physician reviewer.

| Metric | Question Text | Physician Reviewers |
|---|---|---|
| Quality | Overall, this summary is of high quality. | Hospitalists + PCPs |
| Clarity | This summary is appropriately concise and readable. | Hospitalists + PCPs |
| Factuality | The information presented in this summary is accurate and factually correct. | Hospitalists + PCPs |
| Completeness | The summary includes the relevant clinical events that occurred during the patient stay. | Hospitalists + PCPs |
| Ease of Understanding | Overall, I found this summary easy to understand. | PCPs |
| Ease of Verbalization | Based on this summary, it would be easy for me to verbalize the most relevant aspects of the hospitalization to a patient and/or caregiver. | PCPs |
| Ease of Follow-Up | Based on this summary, it would be easy for me to provide follow-up care. | PCPs |

**Supplementary Table S1.** Performance metrics and survey questions assigned to each physician reviewer provider type. Likert scales ranged from 1 to 5, corresponding to strongly disagree, disagree, neutral, agree, and strongly agree.

| **Annotation Error Type** | **Definition** |
|---|---|
| Inaccuracy | Presented information that is factually incorrect or contradicted by the clinical notes. |
| Omission | Missing information that would be detrimental to the patient's post-hospital care if the downstream provider were not informed. |

**Supplementary Table S2.** Definitions for error types included in the instructions for physician reviewer error annotation.

| Metrics | Question Text |
| --- | --- |
| Factuality | Is this incidental finding reported correctly? |
| Clinical Relevance | Should this incidental finding be reported on the discharge summary? |

**Supplementary Table S3.** Metrics and questions for the incidental findings evaluation. Possible answers to these questions were no, unsure and yes.

<table>
<tr><th>Theme</th><th>Sub-themes</th><th>Exemplary quotes</th></tr>
<tr><td>Explanations for higher ratings of LLM-generated summaries over human-authored summaries</td><td>(1) Cognitive burden with summarizing important information chronologically<br><br>(2) Consistency in quality<br><br>(3) Clarity of critical medical information - treatment rationale and unresolved follow-up issues</td><td>Cognitive Burden with summarizing important information chronologically<br><br>“The AI, it was more uniform. Their summaries were just, obviously, I felt more understandable, more succinct, better organized. It told the story. It was easier to read rather than just kind of piecing through it and trying to understand. It was just a lot easier to read and get an idea, an overall idea of the hospital course.” – PCP<br><br>“For me, I think it may be just more of a personal preference, but what helps me understand, of course, the most is the chronological events of, you know, what has happened to lead to me… what is the pathway… that has led to me, and the AI did an exponentially better job at identifying just the cascade of events that led to me as their provider… this is everything that led up until I saw you today.” – Hospitalist<br><br>Consistency in quality<br><br>“For me, I guess the human generated summary, each physician, it was physician dependent, right? So some were better… versus others. Some were quite detailed, others were just maybe 3 paragraphs. Whereas the AI, it was more uniform. Their summaries were… more understandable, more succinct, better organized. It told the story.” – PCP<br><br>“The main things that I noticed between the two, between the hospital summaries and then the AI generated really is the inconsistency between physicians… the redundancy that’s in there… there are so many things that are omitted that are important. It’s just a lot of copy and pasting. Whereas the AI this time, it was concise. There was no redundancy…” – Hospitalist<br><br>Clarity of critical medical information<br><br>“The medication changes were so clear (in LLM-generated summaries). And to me, that’s one of the most important, like, why did you stop this? Why did you add this?" – PCP<br><br>"I also liked one of the AI summaries pulled up, like why a BP med was held where I didn’t get that in the human summary. I probably could have surmised it when I see the patient, like, okay, maybe your blood pressure was low and we’ll reassess it here" – PCP<br><br>“What I’ve loved is that… very commonly there are results that are still pending that kind of, you know, fall through the cracks and anything that was still pending was acknowledged and had a comment on who is appropriate to follow up with these results. … That is something that was</td></tr>
</table>

| | | |
|---|---|---|
| | | essentially not found in any of the… hospitalist-written summaries" – Hospitalist |
| Reasons underlying different ratings of LLM-generated summaries between PCP and hospitalist providers | (1) Differences in perception of error harm potential<br><br>(2) Differences in risk tolerance by care setting<br><br>(3) Differences in workflow | **Differences in perception of error harm likelihood**<br><br>"Yeah, I think probably because we've experienced, you know, somebody sent home on 2 anticoagulants, you know, blood thinners, you know, excessive psychiatric medications, too much diuretics. We see what happens. And often we're the ones that have to deal with the complications" – PCP<br><br>"So my perspective is that we look more at a long-term view of the patient, where they're only with the patient for a short amount of time. So I think from their perspective, being in the hospital, they're looking maybe at low blood pressure or change in vital signs, any, you know, any harm that would happen that they would have to acutely intervene during the hospital admission, whereas we would be dealing as a primary care physician with, you know, long-term consequences of these medicines that should be discontinued, physical and mental health wise." – PCP<br><br>**Differences in risk tolerance by care setting**<br><br>"(PCPs) throw the dice every day… So they probably have seen a lot more stuff than I've seen..So, like, they've seen a lot more of like bad stuff in non-controlled settings." – Hospitalist<br><br>**Differences in workflow**<br><br>"In my practice, I read the discharge summary, the medication reconciliation in there, and then I more often than not don't trust it and go back and I look at vital signs, labs, micro and imaging from the hospitalization, go over it myself." – PCP<br><br>"Especially with these patients that had longer hospitalizations, if a colleague wasn't keeping up an accurate hospital course throughout the notes and then you took over, it is harder to do a very thorough discharge summary and make sure that you're not missing any of those little incidental things." – Hospitalist |

| | | |
|---|---|---|
| LLM identification and reporting of incidental findings | (1) Differential preferences<br><br>(2) Human oversight as non-negotiable | **Differential preferences**<br><br>"[AI] brings it to our attention, which is very important. It doesn't get lost. And then it's our discretion to correlate and see if this needs to be addressed." – Hospitalist<br><br>"As a hospitalist, I am covering my own bases, right? So I'm calling out everything [that could be an incidental finding] and then I let [the PCP] decide which ones he wants to follow or not." – Hospitalist<br><br>"Sometimes they find breast masses, just these incidental findings that need to be followed. And obviously mammograms are not done in the inpatient setting. So those things, I do get every report and I look at them for sure." – PCP<br><br>"I mean, yeah, I don't know (the feelings that AI marks everything). That's a question of like, how much follow up of fatty liver do we need, right? When 30% of the population has fatty, you know, there's, there's, there is always, I don't know, I actually had the opposite reaction where I thought it picked up pretty much everything. Maybe that was just the 10 that I looked at from an incidental finding perspective." – PCP<br><br>**Human oversight as non-negotiable**<br><br>"We're not making a case of AI rooting us out, right? It's just that it's this is the administrative efficiency part of it, right? This still needs to be read by us. That's that should be a disclaimer." – Hospitalist<br><br>"It still requires a human to review it. So I think that kind of helps diminish that issue. I think that it would be very helpful to adopt because you still do need that human lens to make sure that there's accuracy." – Hospitalist<br><br>"I think I'd have to continue to just verify that it is correct. And then maybe after several cases where I determine it is correct, then I would trust it. But right from the bat, I would not. I would definitely have to verify that myself with several patients to make sure it is accurate. " – PCP |
| Implementation considerations for LLM-generated summaries | (1) Human oversight<br><br>(2) Earning trust through repeated verification | **Human oversight** |

| | | |
|---|---|---|
| | (3) Concerns for clinician agency | “No matter what information is pre-populated or anything, it’s still the responsibility of the physician to make sure it’s correct.” – Hospitalist<br><br>“I think it requires the same review that when I do a AI scribe, you know, like I don't just let the AI scribe spit out my note, you know, I have to review it. I change things, it miss things, I reorganize things. But it does, this was this noted in that article I sent out earlier, just this like cognitive offloading. There's so many things that we have. have to keep track of and the idea that that facilitates that. So it doesn't abdicate my responsibility or my to still use my medical intelligence in this process, but I think it speeds it up a little bit. ” – PCP<br><br>**Earning trust through repeated verification**<br><br>"I think I would have to have more experience with AI. I would have to continue my current practice because this is how I do it. And I think I'd have to continue to just verify that it is correct. And then maybe after several cases where I determine it is correct, then I would trust it." – PCP<br><br>“I think (LLM-generated summaries) should decrease the amount of work, but, you know, with the caveat that you got to be careful and you got to trust but confirm. I think that's the term. Trust but verify.” –Hospitalist<br><br>**Concerns for clinician agency**<br><br>“All AI in some sense is going to give up our agency for things. I'm not against technology. I am a proponent of technology. I am just, you know, fearful of the, of not using muscle memory because you get weak then.” – Hospitalist<br><br>"Overall, I am afraid of AI summaries because while it makes your life easier, I guarantee you no one's going back and fact checking or reading everything to make sure that it's fine. It's just going to be, you know, okay, great, it's done and give it to [the PCP]. And [the PCP] still has to do the review unless he totally trusts the AI as the discharging hospitalist does. Because usually when something's handed to you, you're just going to take it as is and you give up your agency to kind of do some of the things that doctors do. So you know, while it's a convenience and it's going to be helpful, I'm afraid that, you know, they're not going to do any QA to make sure that everything is correct." – Hospitalist<br><br>“I think it speeds it up a little bit. with the risk that LLM clearly laid out that, you know, there's a risk that somebody gets lazy and doesn't actually check. ” – PCP |

**Supplementary Table S4:** Supplementary Content from Debrief Sessions

| Length of Stay (Days) | Number of Encounters Overall | Number of Encounters in Sample |
|---|---|---|
| 7 | 69 | 9 |
| 8 | 68 | 9 |
| 9 | 51 | 8 |
| 10 | 40 | 6 |
| 11 | 27 | 4 |
| 12 | 26 | 4 |
| 13 | 22 | 4 |
| 14 | 14 | 2 |
| 15 | 15 | 2 |
| 16 | 11 | 2 |
| 17 | 9 | 2 |
| 18 | 10 | 2 |
| 19 | 9 | 2 |
| 20 | 9 | 2 |
| 21 | 8 | 2 |
| **N** | 385 | 60 |

**Supplementary Table S5.** Distribution of length of stay for overall and study encounters. Overall, N = 385, and for the sample, n = 60.

| **Metric** | **Mean score [SD] difference between LLM-generated vs. human-authored summary (PCPs)** | **Mean score [SD] difference between LLM-generated vs. human-authored summary (Hospitalists)** | **P Value** |
|---|---|---|---|
| Quality | 1.70 [1.33] | 1.40 [1.03] | 0.0865 |
| Conciseness/ Readability | 1.48 [1.28] | 1.05 [1.14] | 0.0235 |
| Factuality | 0.87 [1.10] | 0.92 [0.96] | 0.5907 |
| Completeness | 1.47 [1.38] | 1.07 [0.97] | 0.0219 |

**Supplementary Table S6.** Mean difference in scores between LLM-generated vs. human-authored summaries, stratified by question and reviewer specialty (N = 60).

| Question | Yes | No | Unsure | Percent Yes |
| --- | --- | --- | --- | --- |
| Was this incidental finding reported correctly? | 29 | 1 | 1 | 93.5% |
| Should this incidental finding be reported on the discharge summary? | 27 | 2 | 2 | 87.1% |

**Supplementary Table S7.** Physician reviewer responses to LLM-identified incidental finding questions (N = 31).
